# Identifying Factors Associated with Neonatal Mortality in Sub-Saharan Africa using Machine Learning

**DOI:** 10.1101/2020.10.14.20212225

**Authors:** William Ogallo, Skyler Speakman, Victor Akinwande, Kush R. Varshney, Aisha Walcott-Bryant, Charity Wayua, Komminist Weldemariam, Claire-Helene Mershon, Nosa Orobaton

## Abstract

This study aimed at identifying the factors associated with neonatal mortality. We analyzed the Demographic and Health Survey (DHS) datasets from 10 Sub-Saharan countries. For each survey, we trained machine learning models to identify women who had experienced a neonatal death within the 5 years prior to the survey being administered. We then inspected the models by visualizing the features that were important for each model, and how, on average, changing the values of the features affected the risk of neonatal mortality. We confirmed the known positive correlation between birth frequency and neonatal mortality and identified an unexpected negative correlation between household size and neonatal mortality. We further established that mothers living in smaller households have a higher risk of neonatal mortality compared to mothers living in larger households; and that factors such as the age and gender of the head of the household may influence the association between household size and neonatal mortality.

## Introduction

Improving neonatal outcomes is an important Maternal, Neonatal and Child Health (MNCH) priority for global sustainable development. Despite a global decline in child mortality rates, many countries are not on track to achieving the global targets of ending preventable deaths among newborns and children under 5 years and reducing neonatal mortality to as low as 12 per 1000 live births by the year 2030^1^. Furthermore, the progress towards MNCH-specific targets remains uneven within and across countries^1^, reflected in disparities in access to healthcare services and inequitable allocation of resources for MNCH priorities^2^.

Improved health outcomes rely on interactions of multiple determinants, including socioeconomic factors, health system capacity, and quality of individual care. Coverage of health system interventions and inputs are measured in routine health system data and regular surveys, but they are limited in interpretability for identifying barriers for individual and population-level uptake of high-quality services. Furthermore, due to the limitations of traditional statistical analysis approaches, we are limited to testing identified hypotheses, and it is difficult to generate novel insights from data without the use of machine learning algorithms. However, whereas machine learning algorithms are good at prediction, they are often considered black box models that are selected based on their predictive performance rather than their interpretability and ability to generate actionable insights^3^. Consequently, MNCH stakeholders and policymakers find it difficult to adopt innovative machine learning models for decision-making and intervention planning.

Overall, our research is centered on building machine learning models to characterize the factors associated with poor MNCH health outcomes, and importantly, inspecting these “black box” models to generate actionable insights. We posit that certain subpopulations of mothers and children have an increased risk of shocks (events that impact wellbeing) in their environment, which may be biological or socio-economic, and have limited resilience to respond to these shocks. This combination of risk and lack of resilience may predispose some populations to disproportionately worse health outcomes such as neonatal and maternal mortality (“vulnerable populations”). In this study, we have focused on mothers who are vulnerable to poor neonatal outcomes, namely mortality. Vulnerable populations who have not experienced the same health gains as other groups are the focus of this work, Our overarching goal is to provide MNCH domain experts and stakeholders with capabilities for data- and model-driven decision-making and targeted intervention planning for vulnerable subpopulations and helping to improve equity in access and health outcomes.

In this study, we specifically aim to answer the research question, *which features are associated with neonatal mortality as captured in nationally representative cross-sectional data?* To do so, we analyze the data from the two most recent Demographic and Health Surveys (DHS) from 10 sub-Saharan countries. For each survey, we built an ensemble classifier^4^ using gradient boosting decision trees^5^ to classify the mothers who reported a birth in the 5 years preceding the survey, into those who reported losing one or more children under the age of 28 days and those who did not report losing a child. Subsequently, we inspect each model by visualizing the features in the data that were most important for the model in reaching its conclusion, as well as how changes in the values of the identified features would have impacted neonatal mortality. The primary contribution of this work is to demonstrate how practical machine learning may be used to generate insights about vulnerability to poor health outcomes captured in real-world, nationally representative MNCH survey data. We found that there is significant consistency in MNCH patterns across time and space. We confirmed birth spacing as one of the most important determinants of neonatal mortality and discovered a negative correlation between household size and neonatal mortality that warrants further investigation.

## Related Work

To date, researchers have primarily traced association of neonatal mortality with determinates or particular causes using observational research methods such as cohort and cross-sectional studies along with traditional statistical analyses. For example, Kaguthi et al. conducted a cohort study in western Kenya and used Cox proportional Hazard analysis to identify risk factors neonatal mortality^6^. Similarly, Mengesha et al. conducted a prospective cohort study in northern Ethiopia and subsequently used Kaplan-Meier survival analyses, Log rank test, and Cox-proportional hazard regressions to characterize the survival of neonates and identify the predictors of neonatal mortality in northern Ethiopia^7^. Mekonnen et al. applied a simple linear regression model to examine trends in neonatal mortality rates and a multivariate Cox proportional hazards regression model to examine the factors associated with neonatal mortality in Ethiopian DHS data (2000, 2005, 2011)^8^. Ozodiegwu et al. used logistic regression to estimate the association between maternal obesity and neonatal mortality using DHS data from 34 Sub-Saharan African countries^9^.

Although traditional statistical analyses are well established and effective in addressing certain questions, they are subject to key limitations. For example, domain-specific knowledge is needed for model specification, yet this may impede the discovery of unexpected patterns as the ability of the data to “speak for itself” is limited. Additionally, modeling is often subject to restrictive assumptions such as constraining variables to a linear relationship with the outcome or, in the case of a binary outcome, a linear relationship with the log odds of the outcome. Furthermore, particularly in regression models, modelers typically have to explicitly pre-specify interactions among model variables. Although this makes it easier to identify the relationship between the outcome and individual variables, it limits the ability to discover new interactions between variables that reveal stronger indicators of the outcomes.

Modern machine learning methods address some of the key limitations of traditional statistical approaches and allow for investigators to suspend mental models about how inputs shape outcomes. Interestingly, however, only a few studies have applied such techniques to analyze neonatal mortality. Nesejje and Mwambi compared the performance of using random survival forests to the Cox proportional hazards model and found that the random survival forests can better characterize factors associated with under-5 mortality using DHS data in Uganda^10^. Tesfaye et al. used decision tree classification and rule induction to predict child mortality rates from Ethiopian DHS data^11^, while Kraamwinkel et al. used Bayesian Additive Regression Trees (BART) of conditional average treatment effects to analyze the heterogeneous treatment effect of maternal agency (i.e. education) on severe child under-nutrition in Nigeria^12^.

The gradient boosting classifier applied in this study combines many weak learning models to generate an aggregate model with better performance and improved machine learning results. It is a tried and tested example of ensemble approaches that have shown superior flexibility and accuracy in the generation of state-of-the-art results for prediction and classification tasks^5^. Ensemble methods are appropriately named for their ability to aggregate the outputs of potentially thousands of much simpler models together^4^. This increases their overall accuracy in correctly identifying which records belong to which of the two prediction classes. Furthermore, ensemble methods can model complex nonlinear relationships and do not require the investigator to pre-specify interactions among variables. However, such models provide little knowledge about their internal workings: they are considered to be so-called “black boxes”. To improve explainability, we inspect the models by visualizing feature importance (ranked list of the most important features in an ensemble model) and partial dependence plots (illustration of the relationship between a single feature and an outcome, holding all other features constant). Furthermore, we rely on an interdisciplinary team with content and domain expertise for sensemaking of the insights generated from inspecting our models.

## Methods

In this work, we used cross-sectional data and trained gradient-boosted decision tree models to determine the associations between multiple features and neonatal mortality. Figure 1 illustrates the pipeline used in the study. Although many commonly used machine learning algorithms focus on the ability to predict outcomes, we do not intend to deploy these models for actual prediction purposes. Rather, the training process is to leverage the algorithms’ ability to automatically select and combine features that best demonstrate a correlation with the health outcome of interest.

**Figure 1:**
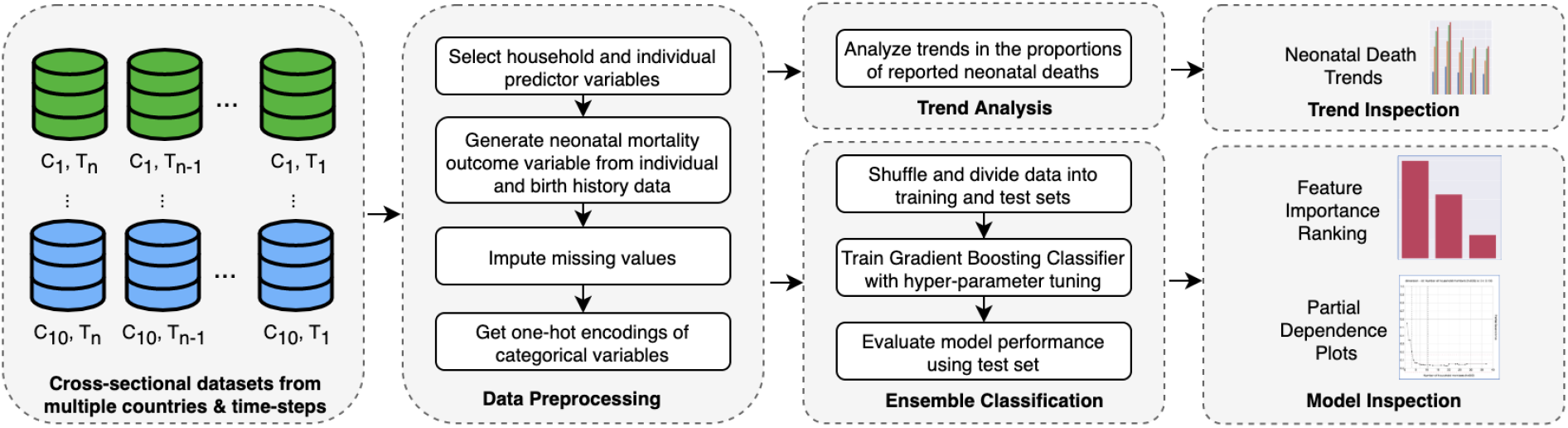
Pipeline for generating neonatal mortality insights from nationally-representative cross-sectional data.

### Datasets

We analyzed DHS data from 10 countries in sub-Saharan Africa, spanning 1998 to 2018 with a total population of 163,180 mothers. These included Burkina Faso (2010, 2003), Democratic Republic of Congo (2013, 2007), Ethiopia (2016, 2011), Ghana (2014, 2008), Kenya (2014, 2008), Nigeria (2018, 2013), Senegal (2017, 2015), Tanzania (2015, 2010), South Africa (2016, 1998), and Zambia (2013, 2007) DHS datasets^13^. DHS data are generated from a series of household surveys conducted in over 90 countries every three to five years. These surveys are nationally representative and are primarily provide data for monitoring and impact assessment of population, health, and nutrition indicators for individual countries, as well as for cross-country comparative analyses^13^. DHS surveys use standard data definitions and data collection procedures across countries and survey phases. In this study, we used household record (HR) files and individual record (IR) files. The HR file contains household data about each surveyed household, while the IR file contains questionnaire and birth history data collected from each woman who is eligible for the DHS survey.

For each survey dataset, we only considered women who reported giving birth at least once in the 5 years before the survey. We defined this unit of analysis as the *mother*. In our analyses, each mother assumes the feature properties of her household in addition to her individual features. We extracted 43 household features (e.g. household size, wealth index, source of water, etc.) and 74 individual features (e.g. age, education level, ethnicity, etc.) giving a total of 117 features. We defined the binary outcomes of interest as neonatal mortality (loss of a newborn ≤28 days old) of a child born in the five years preceding the survey. Mothers who reported the loss of a neonate were assigned to the positive class (1), while mothers who did not report the loss of a neonate were assigned to the negative class (0).

### Mortality Trends

We examined the child mortality trends across DHS surveys in each country categorized as neonatal mortality (death of a live-born child under 28 completed days of life), infant mortality (death of a child before the first birthday), under-2 mortality (death of a child before the second birthday), and under-5 mortality death of a child before the fifth birthday). Note that by definition, these proportions are cumulative. For example, mortality percentages for under-2 represent any child less than 2 years including neonates and infants.

### Modeling and Model Inspection

To investigate the features associated with neonatal mortality, we formulated the problem as a binary classification task. The goal of the modeling task was to train an ensemble gradient boosting classifier^5,14^, such that the aggregated model had the strongest ability to classify individual mothers into one of the two categories of the binary neonatal mortality outcome. For each ensemble model, we used grid search^15^ with stratified 5-fold cross-validation to find the combination of hyperparameter values that maximize the area under the receiver operating characteristic (AUROC) curve of the resulting ensemble model. The model performance results are shown in Table 1. To enable fast training of the models, analyses were done in parallel using a server with 32 core processors and 528 gigabyte memory.

**Table 1:**
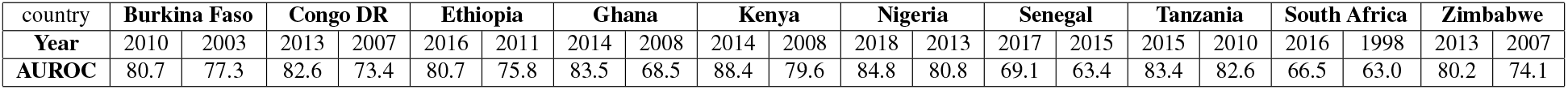
Percent area under receiver operating characteristic (AUROC) curves of the developed ensemble models

Subsequently, we inspected each model using feature importance and partial dependence plots; and visually compared how these vary across time (different DHS survey phases) and space (different countries). *Feature importance* is an average measure of how important a feature is relative to other features used in the ensemble model to predict the outcome. Higher feature importance means that that feature was used to separate one outcome versus the other more often and is a relative measure. *Partial dependence plots*^16^ illustrate how, on average, varying the value of a given feature affects the predicted outcome while holding all other features constant. Such plots can be used to investigate how changes in the values of a feature impact the outcome of interest at the population level.

### Post-Modeling Descriptive Analyses

A majority of our models suggested that *number of births in the last 5 years* before the analyzed survey and *number of household members (household size)* were associated with neonatal mortality (see results section). Whereas it is well-known that birth spacing is associated with neonatal mortality, the association between household size and neonatal mortality was a surprise finding. Accordingly, we investigated this phenomenon further using descriptive analyses. Here, we first binarized the continuous DHS variable for household size (DHS variable hv009) using standard definitions for small (≤ 5 members) vs. large (>5 members) households^17^. We then compared the odds of neonatal mortality in small vs. large households. Furthermore, for each analyzed survey, we compared the odds of having a female head of the household (DHS variable hv219) in small vs. large households. We also evaluated the difference between the mean number of births in the last 5 years per respondent (DHS variable v208) in large vs. small households. Additionally, we evaluated the difference between the mean age of the head of the household (DHS variable hv220), as well as the difference between the mean age of the respondent (DHS variable v012) in large vs. small households.

## Results

### Neonatal Mortality Trends

Figure 2 shows the percentages of mothers who reported the loss of a child (neonate, infant, under-2, or under-5) in that timeframe across multiple DHS phases in the 10 analyzed countries. As a concrete example, for the Nigeria 2018 DHS survey, a total of 41,821 eligible women were interviewed. Of these survey participants, 21,792 reported having given birth in the 5 years before the survey. Of these mothers, 1,239 (5.7%) reported the a neonatal death; 2,019 (9.3%) reported an infant death; 2,726 (12.5%) reported the death of a child under 2 years; and 2,846 (13.0%) reported the death of a child under 5 years. We observe that across all surveys, between 29% and 64% of under-5 deaths are due to neonatal mortality. Interestingly, between 92% and 99% of under-5 deaths occur before the age of 2 years. This is a stratification not usually used in MNCH and provides us with a novel insight into the composition of under 5 deaths. We show that there was an overall decrease in women reporting under-5 deaths in all countries, and the biggest declines began around 2000 in all geographies where data were available. The reduction in women reporting a neonatal death has been much slower, and as a result, neonatal deaths account for the largest proportion of deaths under 5 in the most recent DHS in 8 of the 10 countries. These findings align with published trends in child mortality^18^, and enable MNCH stakeholders to have confidence in the analyses.

**Figure 2:**
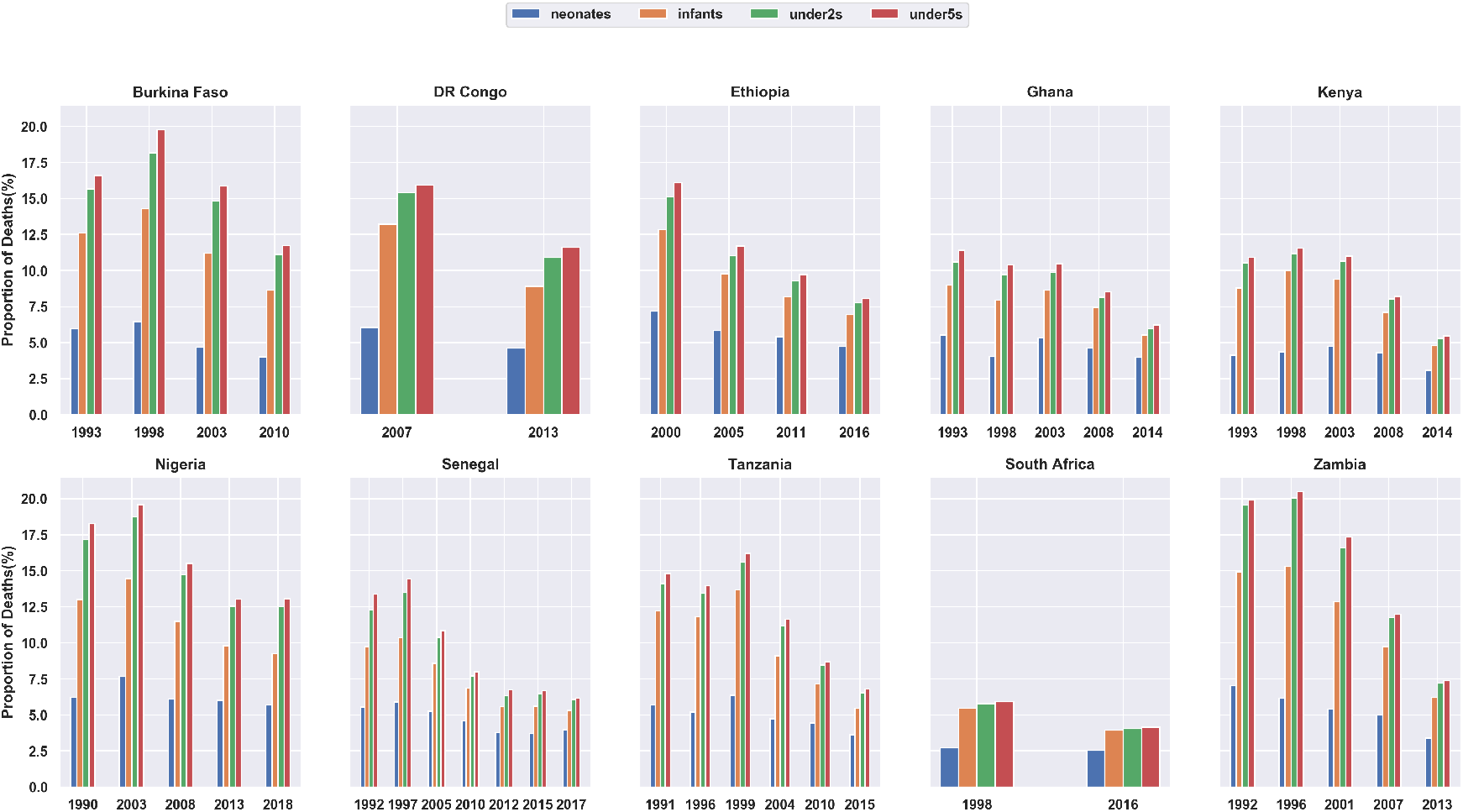
Trends in the proportions women reporting neonatal (blue), infant (orange), under-2 (green) and under-5 (red) deaths in 10 countries. There is an overall decrease in under-5 deaths with smaller decreases in neonatal deaths.

### Factors Associated with Neonatal Mortality

As illustrated in Figure 3, the *number of births in the last 5 years* and the *number of household members* were the “most important” features for predicting whether a mother reported the death of a neonate. Out of the 20 models (10 countries and 2 DHS surveys per country) trained to predict neonatal mortality, the *number of births in the last 5 years* ranked first in all models except Burkina Faso 2003, Tanzania 2015, and Zambia 2007, where the feature ranked second. The *number of household members* ranked first in one model (Zambia 2007), and second in 10 of the 20 models. Together, the two features appeared among the top 5 features in 17 models. Only Ghana 2008, Senegal 2015, and South Africa 2016 did not have the *number of household members* among the top 3 most important features.

**Figure 3:**
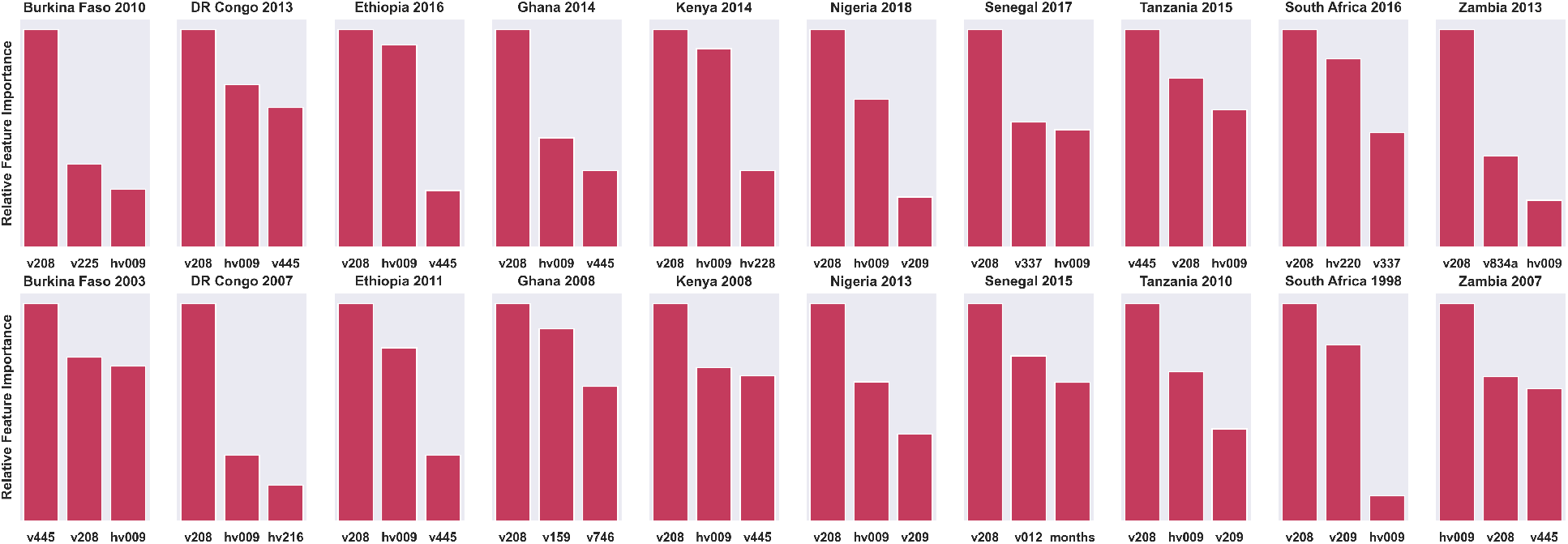
Top 3 features in each of the models for the 2 most recent DHS surveys in 10 countries. *Number of births in the last 5 years (v208)* was present in all models. *Number of household members (hv009)* was present in 17 models. Other frequent features include *body mass index (v445), respondent’s current age (v012), births in past year (v209)*, and *age of head of household (hv220)*.

As shown in Figure 4, the partial dependence plots suggests a positive correlation between the number of births in the last 5 years and the probability of neonatal mortality while holding all other variables constant. This persisted across all the DHS surveys analyzed. Figure 5 illustrates a negative nonlinear correlation between household size and the probability of neonatal mortality, with a large drop between households of size 1 to 4, and a flat line in higher household sizes. This trend persisted in 19 of the 20 models. The only exception was Ghana 2008 which suggests no effect on household size on neonatal mortality for that survey. Interestingly, the feature importance rankings of the neonatal mortality models were also consistent with those of the infant, under-2, and under-5 mortality models.

**Figure 4:**
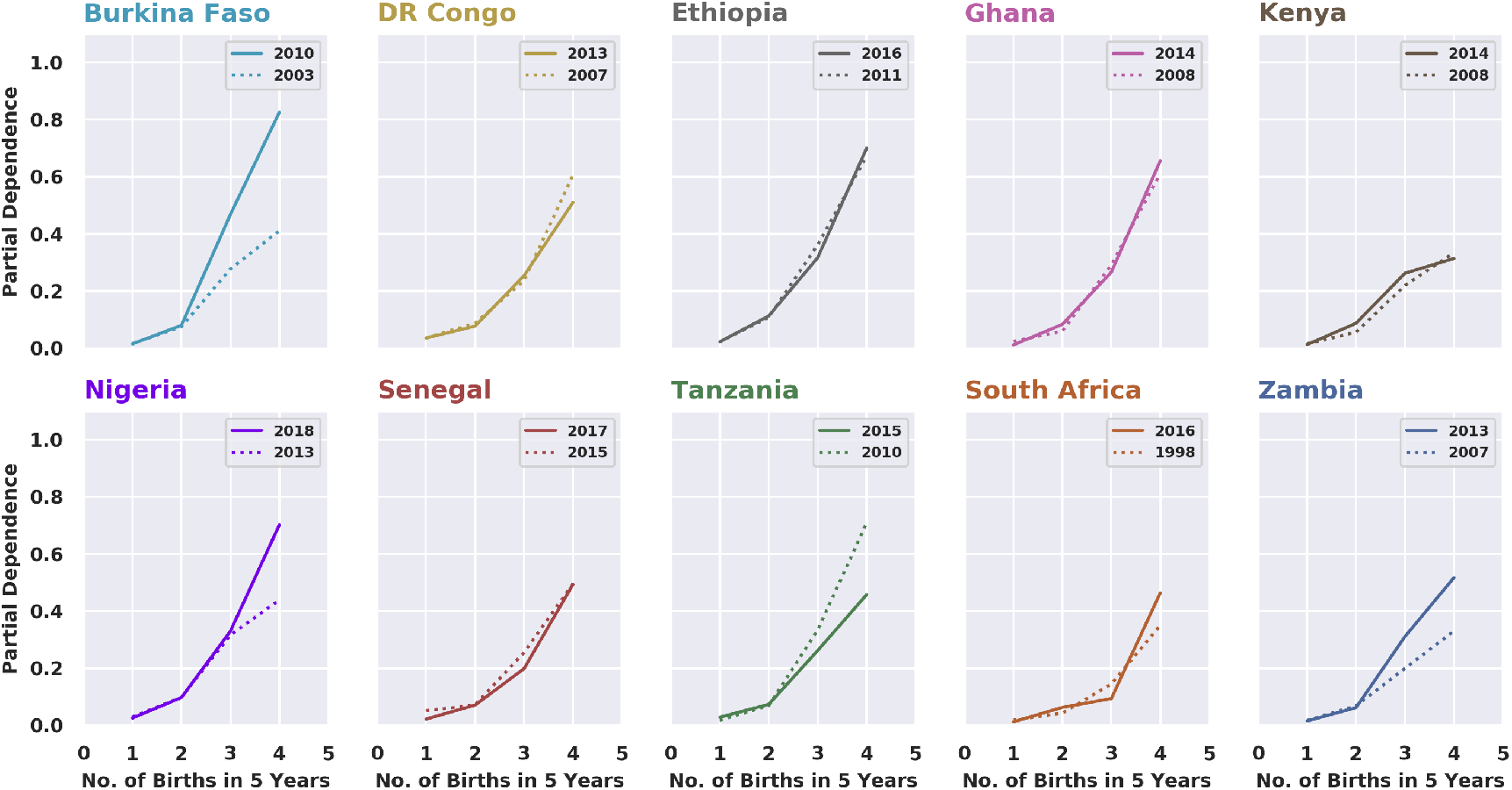
Partial Dependence of Neonatal Mortality on Number of Births in Last 5 years. All models suggested a positive correlation between number of births of the mother and neonatal mortality

**Figure 5:**
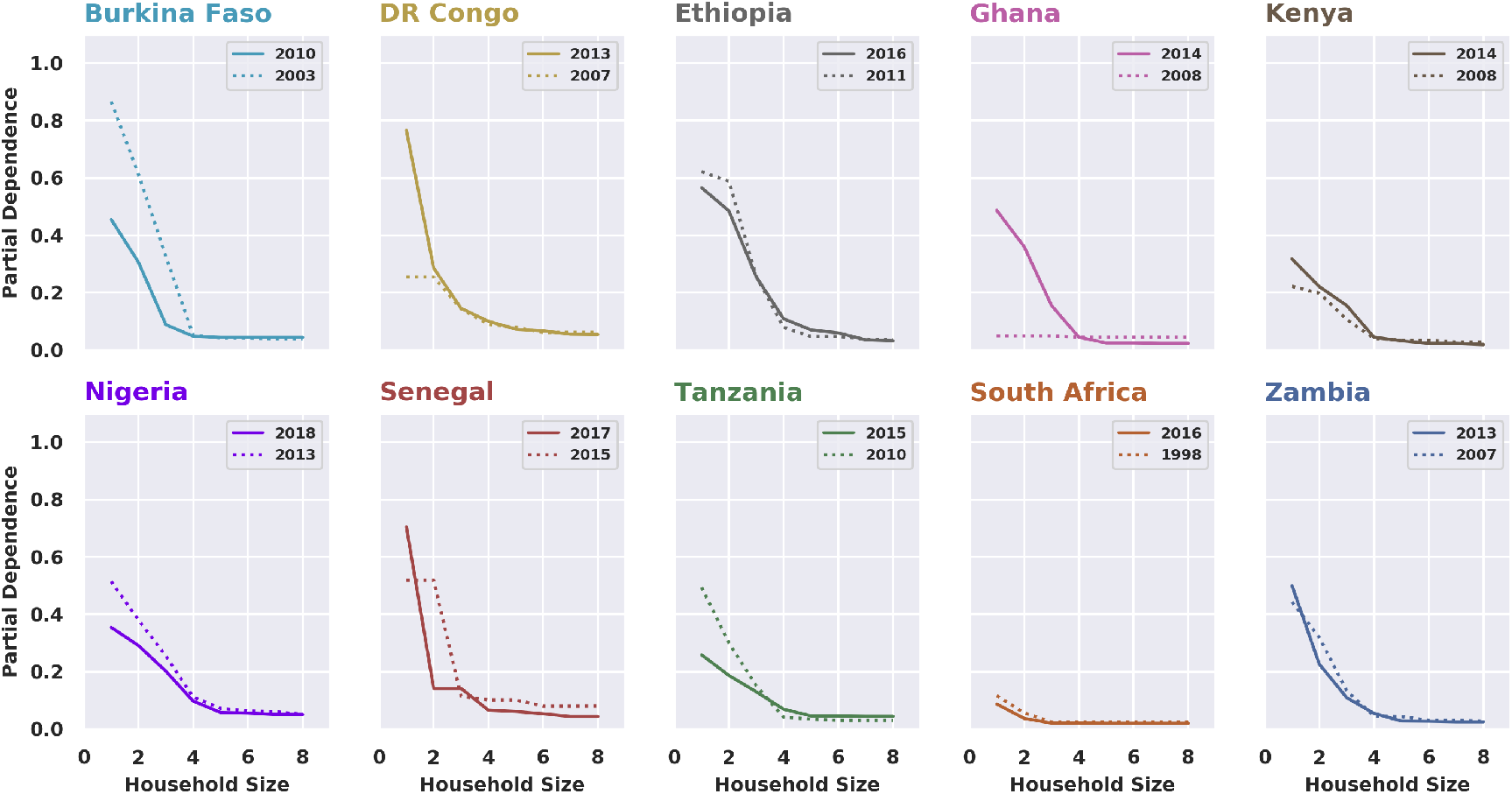
Partial Dependence of Neonatal Mortality on Household Size. All models suggested this negative nonlinear correlation (elbow plot) albeit with varying gradients. The Ghana 2008 model, with a flat line, was the only exception.

Table 2 provides the descriptive statistics of selected variables for women residing in small (≤5 members) vs large (>5 members) households across the two most recent DHS surveys in the 10 countries analyzed. We observe that the proportions of neonatal death were higher in small households compared to large households. This was suggested by our machine learning models and confirmed by our descriptive statistics. Furthermore, in both small and large households, neonatal mortality is high relative to the sustainable development targets for neonatal mortality^1^, and that the magnitude of difference in neonatal mortality in each country is different by the DHS survey year.

**Table 2:** Counts (%), proportions (%), and mean and standard deviation (mean (SD)) of selected variables for mothers living in small (≤5 members) vs large (>5 members) households

| DHS Survey |  | Count (%)<br>Household Size |  | % Neonatal<br>Deaths |  | % Female Head<br>of Household |  | Mean (SD) Births<br>in the last 5 Years |  | Mean (SD) Age<br>of Head of Household |  |
| --- | --- | --- | --- | --- | --- | --- | --- | --- | --- | --- | --- |
| Country | Year | Small | Large | Small | Large | Small | Large | Small | Large | Small | Large |
| Burkina Faso | 2010 | 3734 (36) | 6630 (64) | 4.4 | 3.8 | 11.1 | 4.5 | 1.4 (0.5) | 1.5 (0.6) | 33.9 (10.0) | 46.5 (11.7) |
|  | 2003 | 1850 (25.1) | 5517 (74.9) | 6.4 | 4.1 | 10.8 | 3.2 | 1.4 (0.5) | 1.5 (0.6) | 34.6 (11.0) | 49.0 (13.1) |
| DR Congo | 2013 | 4444 (39.4) | 6849 (60.6) | 5.5 | 4.0 | 27.4 | 16.7 | 1.5 (0.6) | 1.7 (0.7) | 33.0 (10.5) | 43.2 (11.4) |
|  | 2007 | 2161 (39.4) | 3322 (60.6) | 7.5 | 5.1 | 19.9 | 14.7 | 1.6 (0.7) | 1.7 (0.7) | 33.3 (10.4) | 43.5 (11.4) |
| Ethiopia | 2016 | 3548 (49.3) | 3645 (50.7) | 5.6 | 4.0 | 26.7 | 17.6 | 1.3 (0.5) | 1.6 (0.7) | 33.6 (11.1) | 42.7 (11.6) |
|  | 2011 | 3699 (47.6) | 4065 (52.4) | 6.6 | 4.3 | 26.0 | 14.8 | 1.4 (0.6) | 1.6 (0.7) | 32.8 (10.5) | 42.9 (12.1) |
| Ghana | 2014 | 2424 (56.5) | 1870 (43.5) | 4.1 | 3.8 | 32.4 | 15.2 | 1.3 (0.5) | 1.5 (0.6) | 36.4 (11.3) | 45.6 (12.2) |
|  | 2008 | 1172 (54.6) | 975 (45.4) | 4.3 | 5.0 | 35.6 | 18.1 | 1.3 (0.5) | 1.5 (0.6) | 35.9 (11.9) | 46.0 (12.6) |
| Kenya | 2014 | 8167 (54.6) | 6782 (45.4) | 3.5 | 2.6 | 33.9 | 27.6 | 1.3 (0.5) | 1.5 (0.6) | 33.7 (10.4) | 44.0 (12.3) |
|  | 2008 | 2206 (54) | 1876 (46) | 4.7 | 3.8 | 34.9 | 24.4 | 1.4 (0.6) | 1.6 (0.7) | 33.8 (10.9) | 43.9 (12.5) |
| Nigeria | 2018 | 9620 (44.1) | 12172 (55.9) | 6.5 | 5.0 | 15.7 | 6.3 | 1.5 (0.6) | 1.6 (0.7) | 36.6 (11.1) | 45.8 (11.6) |
|  | 2013 | 8486 (42) | 11706 (58) | 7.2 | 5.2 | 17.0 | 7.6 | 1.5 (0.6) | 1.6 (0.7) | 36.0 (11.3) | 45.9 (11.8) |
| Senegal | 2017 | 738 (8.7) | 7748 (91.3) | 5.7 | 3.8 | 39.2 | 22.7 | 1.3 (0.5) | 1.4 (0.6) | 41.3 (12.4) | 53.8 (14.6) |
|  | 2015 | 406 (8.7) | 4275 (91.3) | 3.2 | 3.7 | 39.4 | 22.2 | 1.4 (0.5) | 1.5 (0.6) | 41.1 (12.8) | 52.9 (14.6) |
| Tanzania | 2015 | 2875 (40.8) | 4175 (59.2) | 4.3 | 3.2 | 20.2 | 16.3 | 1.3 (0.5) | 1.5 (0.7) | 35.1 (11.1) | 47.2 (13.0) |
|  | 2010 | 2182 (40.7) | 3176 (59.3) | 5.5 | 3.7 | 20.0 | 16.5 | 1.4 (0.6) | 1.6 (0.7) | 35.2 (10.9) | 46.2 (12.9) |
| South Africa | 2016 | 1711 (56.4) | 1325 (43.6) | 2.8 | 2.2 | 50.1 | 59.8 | 1.1 (0.4) | 1.2 (0.4) | 40.2 (14.7) | 54.9 (15.4) |
|  | 1998 | 1910 (46) | 2238 (54) | 2.8 | 2.7 | 45.6 | 48.2 | 1.2 (0.4) | 1.3 (0.5) | 38.5 (13.3) | 51.9 (14.5) |
| Zambia | 2013 | 3756 (40.2) | 5597 (59.8) | 4.2 | 2.9 | 26.1 | 17.5 | 1.4 (0.5) | 1.5 (0.6) | 33.3 (10.6) | 42.7 (10.8) |
|  | 2007 | 1864 (44.9) | 2284 (55.1) | 6.5 | 3.8 | 24.5 | 16.0 | 1.5 (0.6) | 1.6 (0.6) | 33.3 (10.8) | 42.2 (11.4) |

Figure 6 illustrates the odds of association between household size and neonatal mortality. We observe that the odds of neonatal death across countries and DHS survey timesteps were generally higher in small households compared to large households. For example, for the Nigeria 2018 DHS survey, the odds of neonatal deaths in small households are 1.3 (95% CI: 1.16, 1.46) the odds of neonatal death in large households. Similarly, for the Ethiopia 2016 DHS survey, the odds of neonatal deaths in small households are 1.4 (95% CI: 1.12, 1.74) the odds of neonatal death in large households. Furthermore, given the most recent DHS survey in each analyzed country, only South Africa 2016 (OR = 1.3, 95% CI: 0.82, 2.07), Ghana 2014 1.1 (95% CI: 0.81, 1.5), Burkina Faso 2010 (OR = 1.2, 95% CI: 0.98, 1.47) had odds ratios that were not statistically significant.

**Figure 6:**
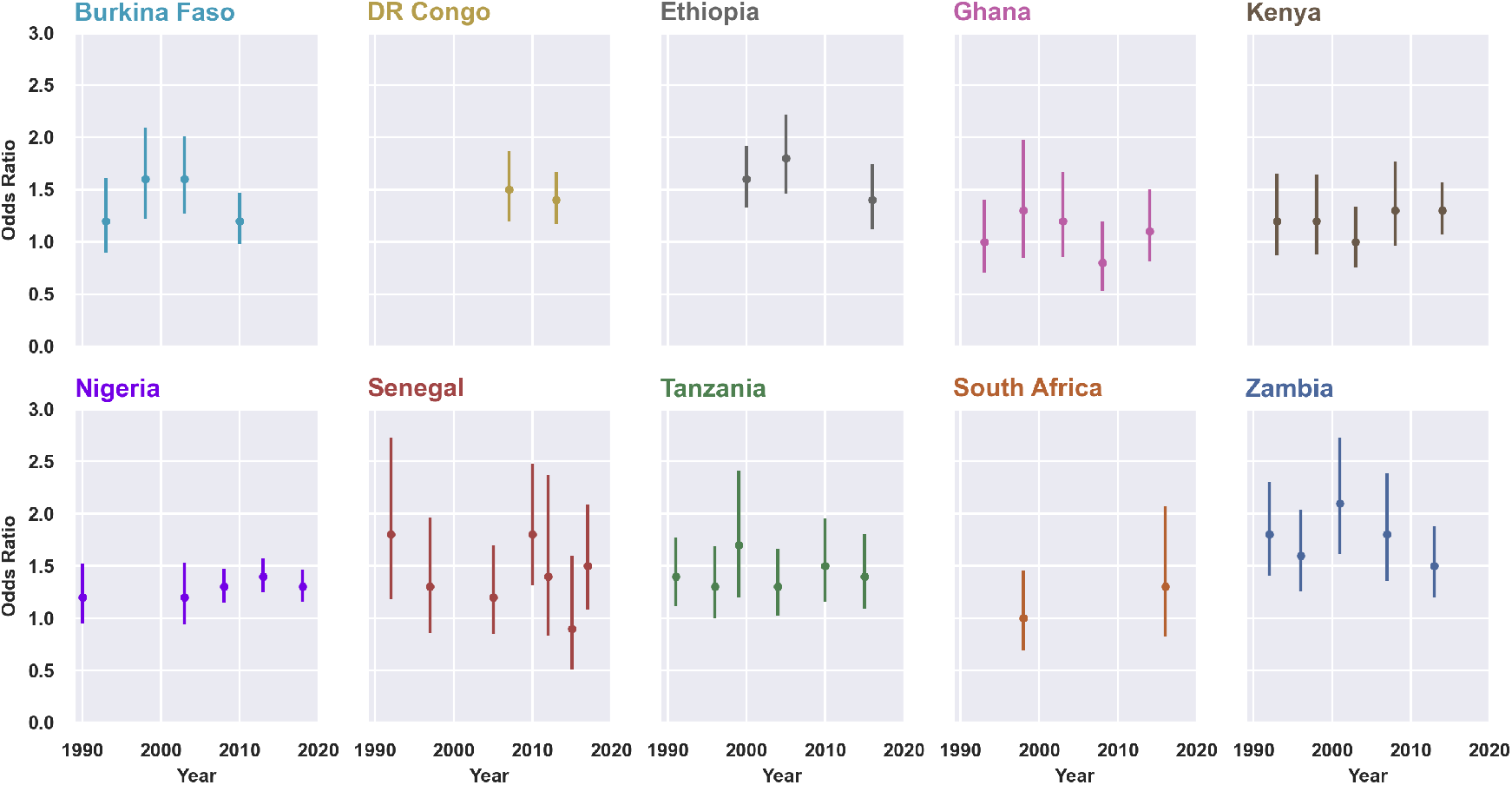
Crude odds ratio (and 95% Confidence Interverals (CIs)) of neonatal death in small vs. large households across survey years in the 10 analyzed countries. Odds ratios with CIs that do not include 1 are statistically significant.

Additionally, we can infer from Table 2 that small households have a higher proportion of female heads of households compared to large households. Furthermore, we established that the crude odds of having a female head of the household were higher in small households compared to large households. This was true for the DHS surveys conducted in 9 out of 10 countries analyzed. Interestingly, the only notable exception is South Africa where the odds of having a female head of the household were high among large households. It is, however, worth noting that the confounders of the observed association between female heads of households and household size were not measured in our study.

We also found that mothers from small households reported fewer births in the past 5 years compared to those from large households for most countries and survey years. These observations are sensible since smaller households may imply younger families. In fact, it is reflected in the observation that small households tended to have younger heads of households. For example, for Nigeria 2018, small households were headed by persons who were 9.2 years (95% CI: 8.9, 9.6) younger than those in large households on average. Other countries and survey timesteps show similar findings that female respondents from smaller households were younger, with the exception of South Africa.

## Discussion

Our study used machine learning to investigate nationally-representative DHS surveys from 10 sub-Saharan countries. Our investigations suggest that in most countries, neonatal deaths accounts for the majority of the loss of children under 5 years and that the percentages of neonatal deaths have historically remained high despite a decrease in under-5 deaths. We found that the number of births in the past 5 years was positively correlated with neonatal mortality, while household size was negatively correlated with neonatal mortality. Furthermore, we established that mothers living in smaller households have a higher risk of neonatal mortality compared to mothers living in larger households. Factors such as the age and gender of the head of the household appear to influence the association between household size and neonatal mortality. It is worth noting that these findings are not causal relationships and that additional work is needed to fully characterize household size as a determinant of neonatal mortality.

The positive correlation between the reported number of births and neonatal mortality reflected in our results confirms the previously known observation about birth spacing as a key determinant of neonatal mortality^19^. Based on evidence from several studies, the World Health Organization recommends waiting at least 2 to 3 years between pregnancies to reduce the likelihood of adverse maternal and neonatal outcomes^20^. In this study, using machine learning, we confirmed that birth spacing is plausibly one of the single most important factors associated with neonatal mortality while accounting for heterogeneity due to other variables. While this finding validates the use of a machine learning approach, it is worth noting that predictors of neonatal mortality are multifactorial. For example, neonatal mortality has been associated with several factors such as short birth intervals^8^, early pregnancy^8^, maternal obesity^9^, maternal education^21^, maternal empowerment (e.g. through family planning choices)^21^, and partner controlling behavior and violence^22^. Furthermore, neonatal mortality is influenced by variability in antenatal care^23^, as well as the location of delivery (e.g. home vs. health facility), skilled attendance at birth, and infections during delivery^21,24^. Although most of these factors were picked by our models, they appeared to be less important for predicting neonatal mortality.

Interestingly, the inverse correlation between household size and neonatal mortality identified in this study was an unexpected finding. There is a dearth of literature on this subject, but it is plausible that household size is a surrogate measure of the capacity of a household to support its mothers and their newborns through, for example, receiving advice from more educated and experienced household members^25^. However, such interpretations must be taken with caution since understanding the correlation between household size and neonatal mortality requires further work to characterize the phenomenon, such as a child’s death moving a household from large to small. Additionally, the household size is measured at the time of the survey as opposed to the time of the death, and may not necessarily reflect the household composition at that time.

To the best of our knowledge, our study is one of the first to apply an ensemble machine learning approach across multiple sub-Saharan DHS surveys. Traditionally, DHS data is primarily designed for generating population-level aggregated statistics such as maternal mortality rates, and the secondary use of DHS data for insight generation is associated with several limitations. For example, since the surveys are self-reported, reporting and recall bias is highly likely for retrospective data relying on the memory of past events by the participants, although less so for events like mortality than for questions like health access. Furthermore, while DHS collects household and individual data, they do not sufficiently collect data on healthcare access and healthcare-seeking behaviors beyond basics like facility delivery and access to antenatal care that would affect outcomes such as neonatal mortality. Additionally, we generated neonatal mortality data directly from the birth histories within each analyzed DHS survey, and this approach may suffer from errors in the data due to the omission of information about the deceased neonates.

A subtle limitation of machine learning models is that they will identify and exploit any pattern made available to them – even if that pattern would be considered ‘cheating’ or ‘obvious’ by a human. This is sometimes referred to as label leakage. We uncovered one such example of label leakage and we acknowledge that there may be others. In the uncovered example, label leakage involved two survey questions that the model learned to combine together. These questions were *How many births in the past 5 years?* and *‘How many children do you have?* If a mother stated that she has 1 child but reported 2 births in the past 5 years, simple subtraction could conclude that one of her children has died. Due to the flexibility of the ensemble models, this type of interaction between variables would be found and used to create a technically more accurate model. However, these are not the type of interactions we wish to discover. To correct for this ‘leaking’ we removed the *How many children do you have?* question from the analysis.

## Conclusion

Despite an overall decline in under-5 deaths over time, the reduction in neonatal deaths has been much lower, and as a result, neonatal deaths account for the largest proportion of child mortality in most countries. Importantly, there is significant consistency in neonatal mortality patterns across time and space. We confirmed birth spacing as one of the most important determinants of neonatal mortality and discovered the inverse relationship between household size and the risk of neonatal mortality that warrants further investigation. As avenues for future work, we will apply other machine learning algorithms to verify findings generated by our ensemble approach. We will use individual conditional expectation (ICE) plots^26^ to visualize and investigate the dependence of neonatal mortality predictions on specific features for individual subjects separately and in comparison to the overall partial dependence plots. Furthermore, we will develop functionalities that enable stakeholders to determine accurate representations of vulnerable subpopulations by identifying and visualizing the ICE plots of subgroups of individuals that show the highest change in prediction (risk of outcome) as the values of a given feature increases or decreases. This will involve developing anomalous pattern detection techniques^27^ for efficiently scanning the exponential number of subsets in a dataset to generate insights about specific subpopulations that are anomalous. Our work demonstrates the practical application of machine learning for generating insights through the inspection of black box models, and the applicability of using machine learning techniques to generate novel insights and alternative hypotheses about phenomena captured in population-level health data.

## Data Availability

This study analyzed the publicly available Demographic and Health Survey (DHS) Data. We applied for and received explicit approval from ICF to access and analyze DHS data from the 10 countries studied: Burkina Faso, Congo Democratic Republic, Ethiopia, Ghana, Kenya, Nigeria, Senegal, South Africa, Tanzania, and Zambia. The approval was received through a letter from the DHS Program's Data Archivist addressed to William Ogallo

https://dhsprogram.com/Data/

## Acknowledgements

This work is funded by Bill & Melinda Gates Foundation, investment ID 52720.

## Notes

### Competing Interest Statement

The authors have declared no competing interest.

### Funding Statement

This study was funded by Bill & Melinda Gates Foundation, investment ID 52720.

### Author Declarations

The protocols, procedures, and questionnaires used to conduct the Demographic and Health Surveys analyzed in this study were reviewed and approved by ICF International's Institutional Review Board (IRB) and host country IRBs for country-specific protocols. The ICF IRB ensures that the survey complies with the U.S. Department of Health and Human Services regulations for the protection of human subjects (45 CFR 46). Host country IRBs ensure that country-specific surveys comply with local laws and norms. The DHS program sought informed consent from all respondents of the DHS surveys before each interview. The informed consent statements provide details regarding the study and emphasized that each respondent's participation was voluntary, the respondent may refuse to answer any question, and the respondent may terminate participation at any time. All respondents's identities and information were kept strictly confidential and all the data were de-identified before being made publicly available.

